# Participant characteristics and exclusion from trials: a meta-analysis of individual participant-level data from phase 3/4 industry-funded trials in chronic medical conditions

**DOI:** 10.1101/2023.04.14.23288549

**Authors:** Jennifer S Lees, Jamie Crowther, Peter Hanlon, Elaine Butterly, Sarah H Wild, Frances S Mair, Bruce Guthrie, Katie Gillies, Sofia Dias, Nicky J Welton, Srinivasa Vittal Katikireddi, David A McAllister

## Abstract

**Objectives:** Trials often do not represent their target populations, threatening external validity. The aim was to assess whether age, sex, comorbidity count and/or race/ethnicity are associated with likelihood of screen failure (i.e., failure to be randomised to the trial for any reason) among potential trial participants.

**Design:** Bayesian meta-analysis of individual participant-level data (IPD).

**Setting:** Industry-funded phase 3/4 trials in chronic medical conditions. Participants were identified as “randomised” or “screen failure” using trial IPD.

**Participants:** Data were available for 52 trials involving 72,178 screened individuals of whom 24,733 (34%) failed screening.

**Main outcome measures:** For each trial, logistic regression models were constructed to assess likelihood of screen failure, regressed on age (per 10-year increment), sex (male versus female), comorbidity count (per one additional comorbidity) and race/ethnicity. Trial-level analyses were combined in Bayesian hierarchical models with pooling across condition.

**Results:** In age- and sex-adjusted models, neither age nor sex was associated with increased odds of screen failure, though weak associations were detected after additionally adjusting for comorbidity (age, per 10-year increment: odds ratio [OR] 1.02; 95% credibility interval [CI] 1.01 to 1.04 and male sex: OR 0.95; 95% CI 0.91 to 1.00). Comorbidity count was weakly associated with screen failure, but in an unexpected direction (OR 0.97 per additional comorbidity, 95% CI 0.94 to 1.00, adjusted for age and sex). Those who self-reported as Black were slightly more likely to fail screening (OR 1.04; 95% CI 0.99 to 1.09); an effect which persisted after adjustment for age, sex and comorbidity count (OR 1.05; 95% CI 0.98 to 1.12).

**Conclusions:** Age, sex, comorbidity count and Black race/ethnicity were not strongly associated with increased likelihood of screen failure. Proportionate increases in screening these underserved populations may improve representation in trials.

**Trial registration:** Relevant trials in chronic medical conditions were identified according to pre-specified criteria (PROSPERO CRD42018048202).

## Background

Women, older people, people with multi-morbidity and those from ethnic minorities (“underserved populations”) are inadequately represented in trials^1–6^. This systematic under-representation undermines the generalisability of trial findings and confidence in the selection of optimal treatment strategies for these groups^7–11^. Furthermore, this under-representation poses ethical issues which can undermine broader public confidence in health research. Despite commitments from funders, journal editors, trialists and policymakers to improve the recruitment and retention of people from underserved populations^2^, there has not been any evident change over the last decade^1,4,5,12–14^.

To become a trial participant, individuals undergo two rounds of selection – an invitation to screening phase and a screening phase proper (Figure 1). In the invitation phase, individuals receive and accept an invitation to attend screening. Such invitees are identified from clinical encounters (or occasionally from patient registers), usually by members of healthcare staff rather than the research trial team. In the screening phase, trial staff apply formal inclusion and exclusion criteria (based on demographic and clinical characteristics) to individuals, and eligible people are invited to participate. The number or characteristics of individuals invited and screened is not a reporting requirement of clinical trial registries such as ClinicalTrials.gov, nor are these items in the influential Consolidated Standards of Reporting Trials (CONSORT) checklist for trial publications^15^. As such, the contribution of invitation and/or screening related factors to underrepresentation is not well described. This is an important gap as it is remains unclear whether changing trial eligibility criteria – in line with recommendations from organisations such as the United States Food and Drug Administration (FDA) – are likely to improve representation.

**Figure 1.**
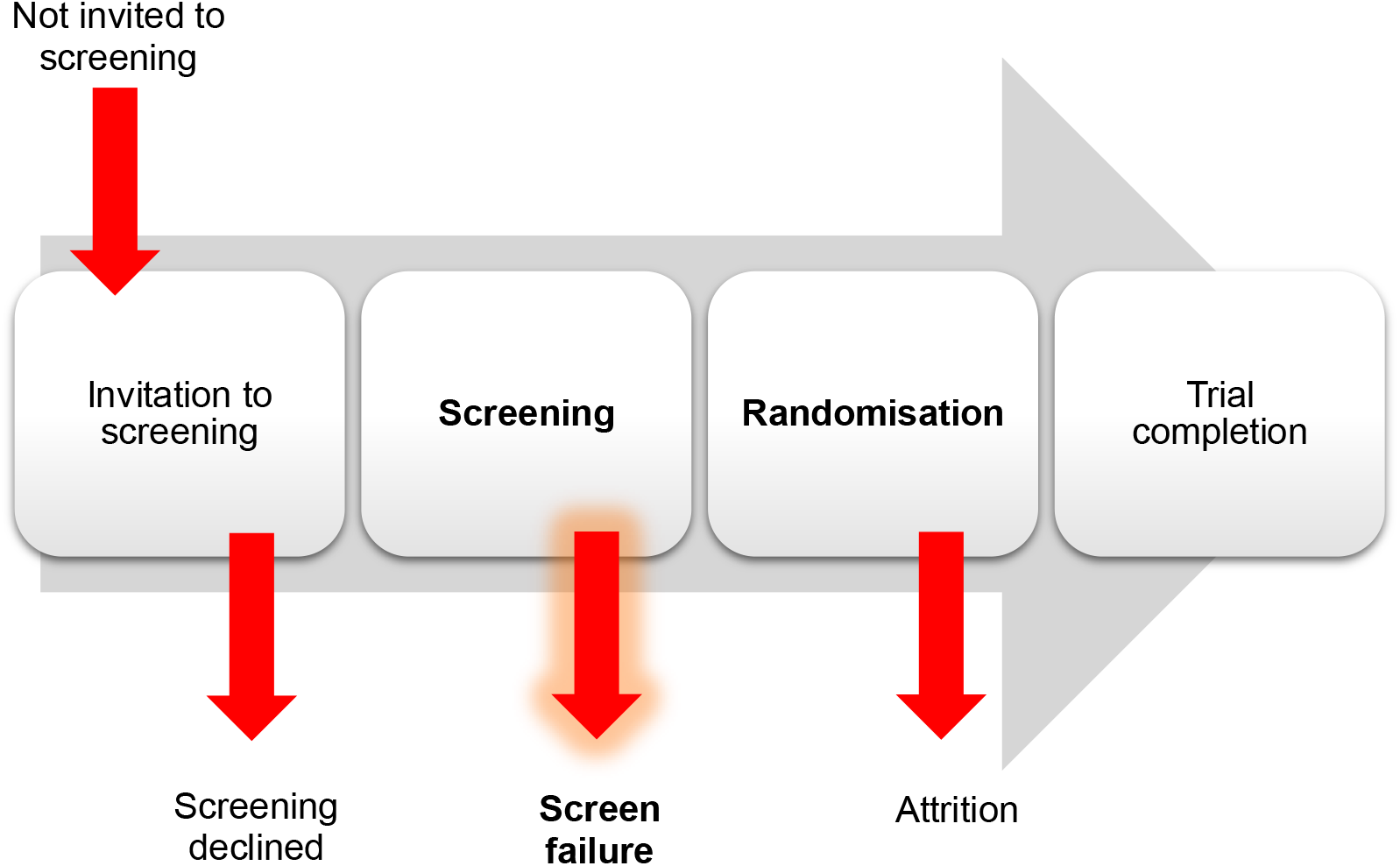
Barriers to trial completion.

We have previously studied age, sex and comorbidity in 116 phase 3/4 industry-funded trials for which we had access to individual participant-level data (IPD). We found that trial participants were younger and had lower comorbidity counts than members of the community with the same index condition^6^. For a subset of these trials, we have access to data on individuals screened for participation. Therefore, we now examine whether age, sex, comorbidity counts, and self-reported race/ethnicity, predict failure to progress to randomisation among individuals who have undergone trial screening.

## Methods

### Study design

This was a Bayesian meta-analysis of IPD from industry-funded phase 3 or 4 trials in chronic medical conditions. We explore whether individual demographic and clinical characteristics are associated with failing trial screening for any reason.

### Data sources and participants

Available IPD were obtained from Phase 3 or 4 trials contained within the Vivli trial repository (https://vivli.org). Appropriate trials for inclusion were identified according to pre-specified criteria (PROSPERO CRD42018048202)^6^. In brief, we included trials conducted in chronic medical conditions that are managed pharmacologically (but excluding trials in cancer, infections, psychiatric and developmental disorders)^6^.

Participants were identified as “randomised” or “screen failure” from within the trial IPD (Figure 1). Age, sex, comorbidity count and race/ethnicity were extracted where available for randomised participants and screen failures.

As previously described^6,16,17^, comorbidities were defined using concomitant medications and pre-specified medical history-based definition (MedDRA codes) for cardiovascular disease, chronic pain, arthritis, affective disorders, acid-related disorders, asthma/chronic obstructive pulmonary disease, diabetes mellitus, osteoporosis, thyroid disease, thromboembolic disease, inflammatory conditions, benign prostatic hyperplasia, gout, glaucoma, urinary incontinence, erectile dysfunction, psychotic disorders, epilepsy, migraine, parkinsonism and dementia^6^. Individuals were considered to have a comorbidity if they had evidence of this comorbidity from either concomitant medications or medical history-based definition (or both). A comorbidity count was calculated as the sum of the number of comorbidities at baseline (excluding the index condition).

### Outcome

The outcome of interest was “screen failure”, defined as a failure to be randomised to a treatment group for any reason after entering the screening process.

### Statistical analysis

Participant characteristics for randomised participants and screen failures were calculated for each available IPD trial. These included: age (mean and standard deviation; SD); sex (number and %); comorbidity count (mean and SD); number with 0, 1 and 2 or more comorbidities; race/ethnicity (number and %). Race/ethnicity categories used in this analysis were largely driven by those recorded in the trial IPD, which included White, Black or African descent (“Black”), Asian, American Indian or Alaska Native and Native American or Other Pacific Islander (“Indigenous”) and multiple or other “Other”. Of these the first four were as per the FDA recommendations^18^; the fifth “Other” was formed by collapsing all other categories because of small numbers.

Full details of the modelling have been published previously^16^. In brief, for each trial, logistic regression models were constructed to assess likelihood of screen failure, regressed on age (per 10-year increment), sex (male versus female), comorbidity count (per 1 additional comorbidity) and race/ethnicity.

Coefficients, standard errors and variance/covariance matrices were exported for each model from the Vivli secure environment. The estimates from each trial were then meta-analysed in Bayesian hierarchical models. Each model had a multivariate normal likelihood, where for each trial the exported coefficients supplied a vector of means, and the exported variance-covariance matrices for these coefficients was the covariance matrix of the multivariate normal. In the primary analysis, models were fitted with trial nested within index condition. In secondary analyses, we then explored different structures for the model hierarchy, where trial was nested within both index condition and treatment comparison. For all models, trial was treated as a random effect.

We fitted models with five main sets of covariates: i) age and sex; ii) comorbidity count; iii) race/ethnicity; iv) age, sex and comorbidity count, and; v) age, sex, comorbidity count and race/ethnicity. We fitted additional models with interaction terms for selected two-way and three-way interactions. For sex, female was the reference category, while for race/ethnicity, White was the reference category (as this was the largest group and present in all trials) with dummy (indicator) variables for the remaining levels. In sensitivity analyses, we included only participants with one or more, or two or more comorbidities (other than the index condition).

For each model we report point estimate and 95% credible interval odds ratios for the association between each characteristic (age, sex, comorbidity count and race/ethnicity) and screen failure. These were obtained by exponentiating the posterior distributions and obtaining the mean, 2.5^th^ and 97.5^th^ percentiles. We additionally report between trial, between index condition and between treatment comparison variation for each parameter as the standard deviations. Finally, for model v) we present condition-specific odds ratios.

## Results

### Baseline data

There were 52 trials involving 72,178 screened individuals, of whom 24,733 (34%) failed screening (Table 1). The number of trials included in the sequential models reflected the data availability in the trials. Age and sex data were available for all 52 trials. Comorbidity count data (including for individuals who did not pass screening) were available for 31 trials and race/ethnicity data were available for 45 trials. Both race/ethnicity and comorbidity count data were available for 27 trials.

**Table 1.** Characteristics of randomised participants (Randomised = “Yes”) and screen failures (Randomised = “No”) for included studies. Race/ethnicity categories included White (“White”), Black or African descent (“Black”), Asian (“Asian”), American Indian or Alaska Native and Native American or Other Pacific Islander (“Indigenous”) and Multiple or Other (“Other”).

| Index condition | Randomised | N trials | N participants | Male: n(%) | Age (years) | Trials with comorbidity count | Comorbidity count = 0: n(%) | Comorbidity count = 1: n(%) | Comorbidity count >= 2: n(%) | Trials with ethnicity: n(%) | White | Asian | Black | Other | Indigenous |
| --- | --- | --- | --- | --- | --- | --- | --- | --- | --- | --- | --- | --- | --- | --- | --- |
| Asthma | Yes | 4 | 1625 | 652 (40.1%) | 43 (17) | 0 |  |  |  | 4 (1625) | 1097 (67.5%) | 112 (6.9%) | 134 (8.2%) | 282 (17.4%) | 0 (0%) |
| Asthma | No | 4 | 1460 | 572 (39.2%) | 42 (17) | 0 |  |  |  | 4 (1460) | 1024 (70.1%) | 63 (4.3%) | 219 (15%) | 154 (10.5%) | 0 (0%) |
| BPH | Yes | 4 | 1783 | 1783 (100%) | 64 (8) | 3 (1458) | 475 (32.6%) | 428 (29.4%) | 555 (38.1%) | 3 (1154) | 969 (84%) | 4 (0.3%) | 40 (3.5%) | 5 (0.4%) | 136 (11.8%) |
| BPH | No | 4 | 100 | 100 (100%) | 65 (8) | 3 (84) | 32 (38.1%) | 20 (23.8%) | 32 (38.1%) | 3 (83) | 71 (85.5%) | 1 (1.2%) | 5 (6%) | 0 (0%) | 6 (7.2%) |
| Dementia | Yes | 1 | 580 | 216 (37.2%) | 72 (8) | 1 (580) | 349 (60.2%) | 120 (20.7%) | 111 (19.1%) | 1 (580) | 418 (72.1%) | 143 (24.7%) | 8 (1.4%) | 11 (1.9%) | 0 (0%) |
| Dementia | No | 1 | 58 | 19 (32.8%) | 75 (8) | 1 (58) | 33 (56.9%) | 11 (19%) | 14 (24.1%) | 1 (58) | 42 (72.4%) | 13 (22.4%) | 1 (1.7%) | 2 (3.4%) | 0 (0%) |
| Diabetes | Yes | 12 | 17121 | 10116 (59.1%) | 58 (10) | 8 (6829) | 1512 (22.1%) | 1758 (25.7%) | 3559 (52.1%) | 9 (8545) | 5646 (66.1%) | 1129 (13.2%) | 429 (5%) | 808 (9.5%) | 530 (6.2%) |
| Diabetes | No | 12 | 10568 | 5992 (56.7%) | 59 (11) | 8 (2720) | 1037 (38.1%) | 880 (32.4%) | 803 (29.5%) | 9 (3744) | 1175 (31.4%) | 180 (4.8%) | 217 (5.8%) | 199 (5.3%) | 90 (2.4%) |
| ED | Yes | 1 | 606 | 606 (100%) | 63 (8) | 1 (606) | 235 (38.8%) | 174 (28.7%) | 197 (32.5%) | 1 (606) | 565 (93.2%) | 14 (2.3%) | 23 (3.8%) | 3 (0.5%) | 1 (0.2%) |
| ED | No | 1 | 132 | 132 (100%) | 63 (9) | 1 (132) | 50 (37.9%) | 36 (27.3%) | 46 (34.8%) | 1 (132) | 125 (94.7%) | 2 (1.5%) | 5 (3.8%) | 0 (0%) | 0 (0%) |
| Heart failure | Yes | 1 | 107 | 64 (59.8 %) | 57 (11) | 1 (107) | 26 (24.3%) | 47 (43.9%) | 34 (31.8%) | 1 (107) | 76 (71%) | 15 (14%) | 16 (15%) | 0 (0%) | 0 (0%) |
| Heart failure | No | 1 | 159 | 90 (56.6 %) | 59 (10) | 1 (159) | 62 (39%) | 43 (27%) | 54 (34%) | 1 (159) | 115 (72.3 %) | 17 (10.7 %) | 27 (17%) | 0 (0%) | 0 (0%) |
| Hypertension | Yes | 8 | 5473 | 3181 (58.1 %) | 56 (11) | 7 (5047) | 2767 (54.8%) | 1496 (29.6%) | 784 (15.5%) | 7 (4672) | 3914 (83.8 %) | 225 (4.8%) | 312 (6.7%) | 80 (1.7%) | 0 (0%) |
| Hypertension | No | 8 | 3290 | 1547 (47%) | 56 (11) | 7 (2880) | 1531 (53.2%) | 892 (31%) | 457 (15.9%) | 7 (2527) | 2237 (88.5 %) | 30 (1.2%) | 175 (6.9%) | 34 (1.3%) | 0 (0%) |
| Hypertension, Pulmonary | Yes | 1 | 406 | 88 (21.7 %) | 54 (16) | 0 |  |  |  | 1 (406) | 327 (80.5 %) | 34 (8.4%) | 35 (8.6%) | 2 (0.5%) | 6 (1.5%) |
| Hypertension, Pulmonary | No | 1 | 62 | 15 (24.2 %) |  | 0 |  |  |  | 1 (62) | 48 (77.4 %) | 7 (11.3 %) | 5 (8.1%) | 0 (0%) | 2 (3.2%) |
| Osteoporosis | Yes | 3 | 10976 | 84 (0.8%) | 67 (8) | 1 (2088) | 3 (0.1%) | 565 (27.1%) | 1520 (72.8%) | 3 (10976) | 10419 (94.9 %) | 164 (1.5%) | 60 (0.5%) | 333 (3%) | 0 (0%) |
| Osteoporosis | No | 3 | 4283 | 64 (1.5%) | 67 (8) | 1 (1557) | 42 (2.7%) | 478 (30.7%) | 1037 (66.6%) | 3 (4283) | 4022 (93.9 %) | 38 (0.9%) | 29 (0.7%) | 194 (4.5%) | 0 (0%) |
| Parkinson Disease | Yes | 3 | 1368 | 791 (57.8 %) | 62 (10) | 2 (1057) | 428 (40.5%) | 322 (30.5%) | 307 (29%) | 2 (1057) | 602 (57%) | 455 (43%) | 0 (0%) | 0 (0%) | 0 (0%) |
| Parkinson Disease | No | 3 | 171 | 74 (43.3 %) | 69 (10) | 2 (159) | 58 (36.5%) | 47 (29.6%) | 54 (34%) | 2 (159) | 70 (44%) | 89 (56%) | 0 (0%) | 0 (0%) | 0 (0%) |
| Pulmonary Disease, Chronic Obstructive | Yes | 6 | 4385 | 2870 (65.5 %) | 64 (8) | 5 (3539) | 861 (24.3%) | 1159 (32.7%) | 1519 (42.9%) | 6 (4385) | 4069 (92.8 %) | 144 (3.3%) | 73 (1.7%) | 99 (2.3%) | 0 (0%) |
| Pulmonary Disease, Chronic Obstructive | No | 6 | 1322 | 793 (60%) | 65 (9) | 5 (1027) | 502 (48.9%) | 203 (19.8%) | 322 (31.4%) | 6 (1322) | 1197 (90.5 %) | 44 (3.3%) | 31 (2.3%) | 44 (3.3%) | 0 (0%) |
| Restless Legs Syndrome | Yes | 1 | 331 | 133 (40.2 %) | 57 (12) | 1 (331) | 84 (25.4%) | 110 (33.2%) | 137 (41.4%) | 0 |  |  |  |  |  |
| Restless Legs Syndrome | No | 1 | 166 | 51 (30.7 %) | 56 (15) | 1 (166) | 60 (36.1%) | 53 (31.9%) | 53 (31.9%) | 0 |  |  |  |  |  |
| Rhinitis | Yes | 7 | 2684 | 933 (34.8 %) | 40 (14) | 1 (302) | 203 (67.2%) | 62 (20.5%) | 37 (12.3%) | 7 (2684) | 2057 (76.6 %) | 46 (1.7% ) | 480 (17.9 %) | 101 (3.8% ) | 0 (0%) |
| Rhinitis | No | 7 | 2962 | 1065 (36%) | 40 (16) | 1 (267) | 213 (79.8%) | 37 (13.9%) | 17 (6.4%) | 7 (2962) | 2013 (68%) | 57 (1.9% ) | 803 (27.1 %) | 89 (3%) | 0 (0%) |
| All | Yes | 52 | 47445 | 21517 (45.4 %) | 59 (13) | 31 (21944) | 6943 (31.6%) | 6241 (28.4%) | 8760 (39.9%) | 45 (36797) | 30159 (82%) | 2485 (6.8% ) | 1610 (4.4% ) | 1724 (4.7% ) | 673 (1.8%) |
| All | No | 52 | 24733 | 10514 (42.5 %) | 57 (14) | 31 (9209) | 3620 (39.3%) | 2700 (29.3%) | 2889 (31.4%) | 45 (16951) | 12139 (71.6 %) | 541 (3.2% ) | 1517 (8.9% ) | 716 (4.2% ) | 98 (0.6%) |

### Trial-level factors and screen failure

On visual inspection, there were no identifiable associations between year of trial conduct, trial size or trial phase and proportion of individuals who did not pass screening (Figure 2). Trial-level factors were not explored further in meta-analysis models.

**Figure 2.**
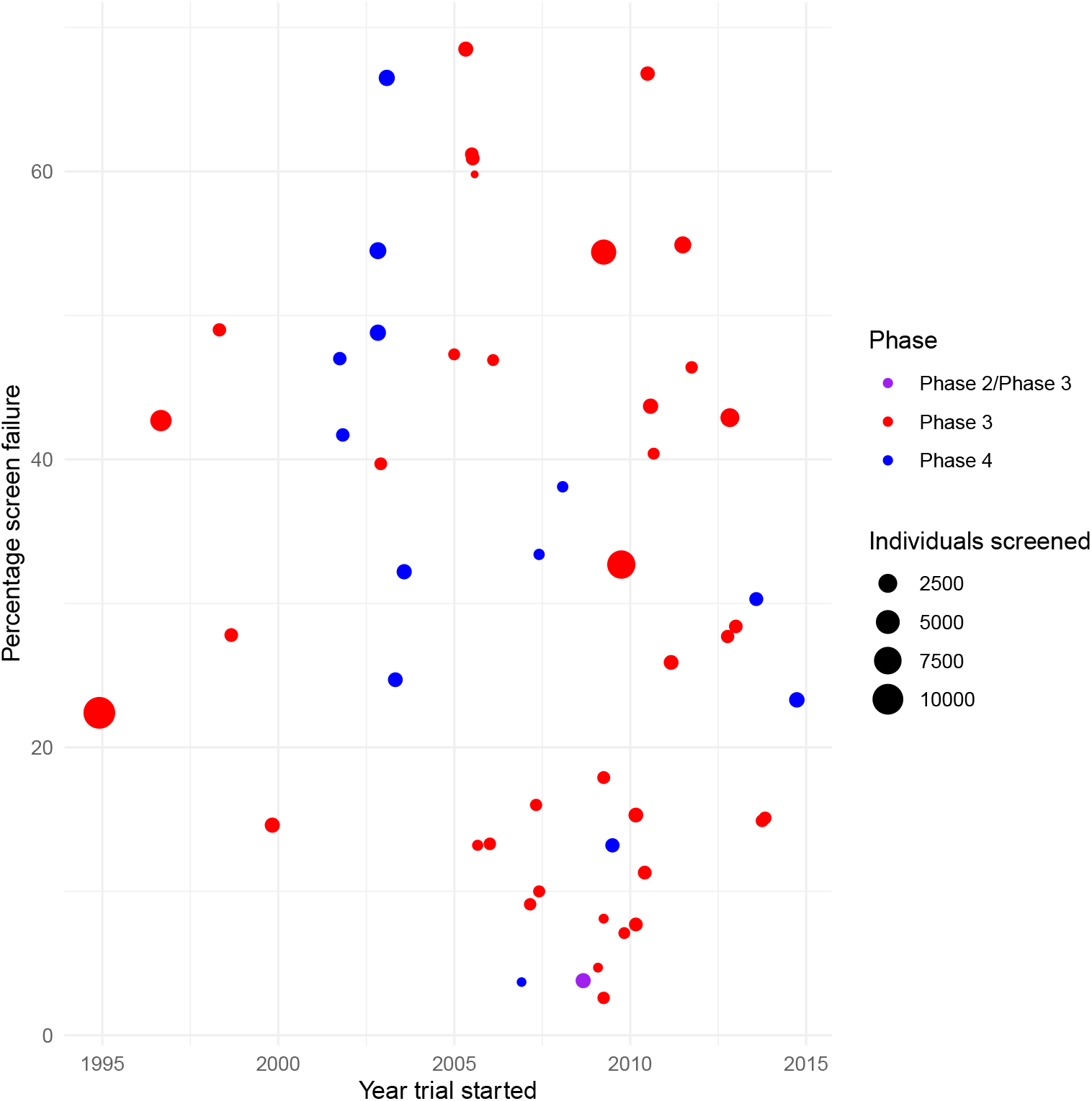
Scatter plot of trial-level factors against percentage of participants who failed screening for any reason.

### Individual participant-level factors associated with screen failure: primary analysis with trial nested within index condition

On modelling age and sex (n=52 trials), neither was associated with increased odds of screen failure (odds ratio [OR] 1.01; 95% CI 0.99 to 1.03 for age per 10-year increment; OR 0.97; 95% CI 0.93 to 1.01 for male versus female sex; Table 2). After additional adjustment for comorbidity count (n=31 trials), there was a weak association between reduced odds of screen failure for older age and for male sex, though the latter just crossed the null (Table 2).

**Table 2.**
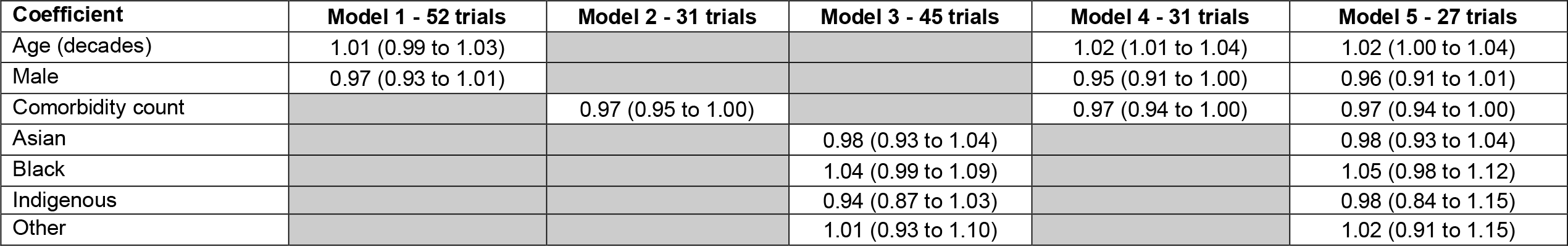
Trial level models for the mean odds ratio (standard error) [95% credible interval] for screen failure. Model 1: adjusted for age and sex; Model 2: comorbidity count only; Model 3: race/ethnicity only; Model 4: adjusted for age, sex and comorbidity count; Model 5: adjusted for age, sex, comorbidity count and race/ethnicity. For all models, trial was nested within index condition.

Comorbidity count was weakly associated with screen failure, but in an unexpected direction (n=31 trials). In a model including solely comorbidity count the OR was 0.93 per additional comorbidity (95% CI 0.87 to 1.00). On additionally adjusting for age and sex (n=31 trials), and age, sex and race/ethnicity (n=27 trials) the odds ratios were similar (Table 2). In sensitivity analyses (restricting analyses to participants with one or more, or two or more comorbidities), the association between comorbidity count and screen failure was considerably weaker, the credible intervals included the null and overall were consistent with no association (OR 0.99; 95% CI 0.97 to 1.02 for both sensitivity analyses).

On modelling race/ethnicity in a univariate analysis (n=45 trials), all the credible intervals included the null (Table 2); however, self-reported Black race/ethnicity appeared to be weakly associated with higher likelihood of failing screening (OR 1.04; 95% CI 0.99 to 1.09). After adjustment for age, sex and comorbidity count (n=27 trials), the point estimate and credible interval was similar (OR 1.05; 95% CI 0.98 to 1.12).

There was no evidence of an interaction between age and sex, sex and comorbidity count, or age, sex and comorbidity count (Supplementary Table S1). On modelling an interaction between male sex and Black race/ethnicity, there was a slightly stronger association in women (OR 1.09; 95% CI 1.00 to 1.19) than men (OR 0.97; 0.75 to 1.19). However, the credible interval for the odds ratio for the interaction included the null (OR 0.90; 0.70 to 1.08) and this comparison should be interpreted circumspectly.

### Variation across trials, index conditions and treatment comparisons: secondary analyses

The associations between individual participant-level factors and screen failure were similar in models where index condition and treatment comparison were ignored, and where trial was nested within both index condition and treatment comparison (Table 3 and Supplementary Table S2). There was no evidence of substantial variation across index conditions or treatment comparisons for associations between age, race/ethnicity and screen failure (Table 3 and Figure 3). There was a clear association between male sex and reduced odds of screen failure in trials in hypertension and chronic obstructive pulmonary disease (Figures 3 and 4). For comorbidity count, although the point estimates were in the same direction (below one) for all the index conditions, the associations were more markedly negative for asthma and for rhinitis, and to a lesser extent, for osteoporosis and diabetes (Figures 3 and 4).

**Table 3.**
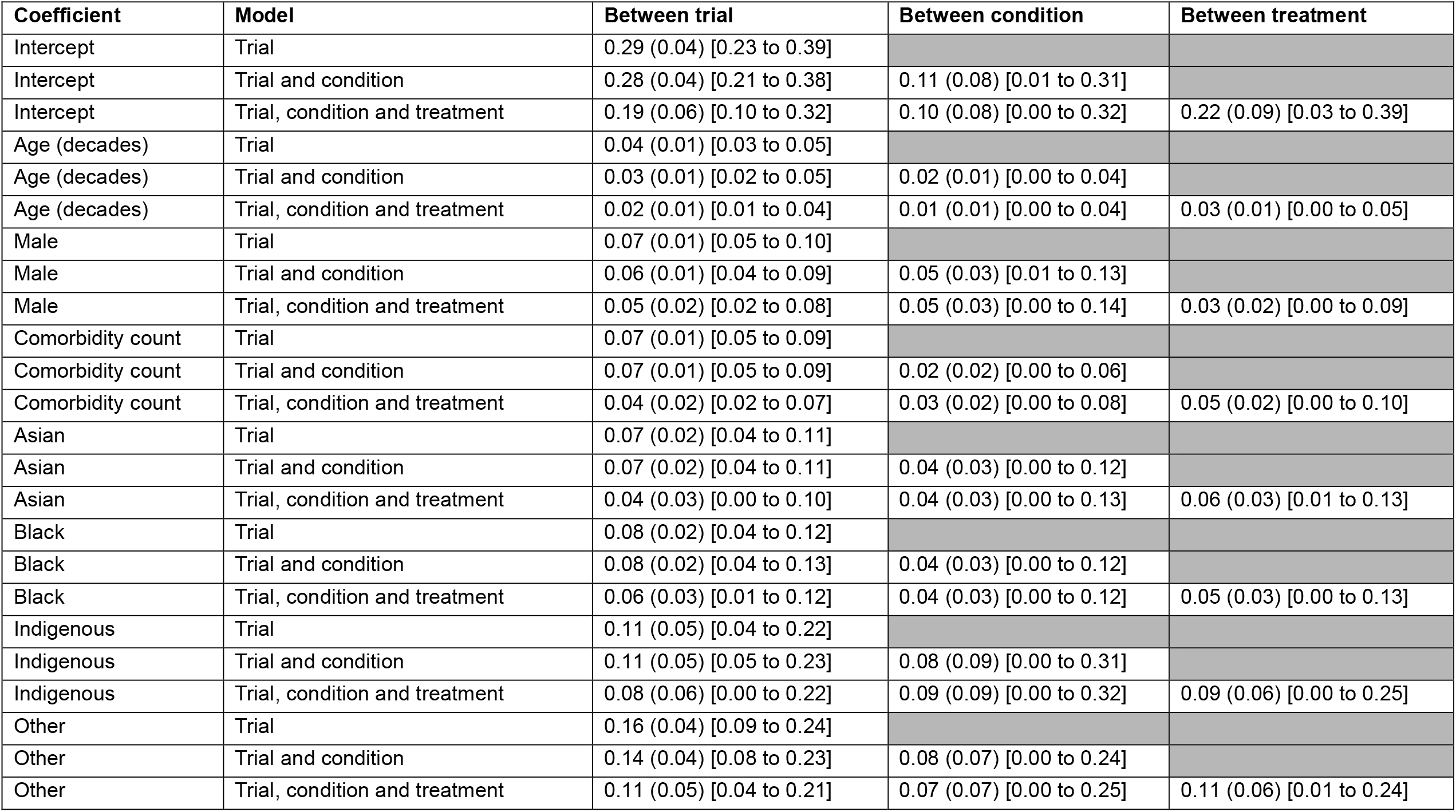
Models for the log odds ratio (standard error) [95% credible interval] for screen failure examining variation in estimates between trial, between condition and between trial and condition. Models, adjusted for age, sex, comorbidity count and race/ethnicity, were conducted at three levels: trial (where condition and treatment were ignored); trial nested within condition; and trial nested within condition and treatment.

| Coefficient | Model | Between trial | Between condition | Between treatment |
| --- | --- | --- | --- | --- |
| Intercept | Trial | 0.29 (0.04) [0.23 to 0.39] |  |  |
| Intercept | Trial and condition | 0.28 (0.04) [0.21 to 0.38] | 0.11 (0.08) [0.01 to 0.31] |  |
| Intercept | Trial, condition and treatment | 0.19 (0.06) [0.10 to 0.32] | 0.10 (0.08) [0.00 to 0.32] | 0.22 (0.09) [0.03 to 0.39] |
| Age (decades) | Trial | 0.04 (0.01) [0.03 to 0.05] |  |  |
| Age (decades) | Trial and condition | 0.03 (0.01) [0.02 to 0.05] | 0.02 (0.01) [0.00 to 0.04] |  |
| Age (decades) | Trial, condition and treatment | 0.02 (0.01) [0.01 to 0.04] | 0.01 (0.01) [0.00 to 0.04] | 0.03 (0.01) [0.00 to 0.05] |
| Male | Trial | 0.07 (0.01) [0.05 to 0.10] |  |  |
| Male | Trial and condition | 0.06 (0.01) [0.04 to 0.09] | 0.05 (0.03) [0.01 to 0.13] |  |
| Male | Trial, condition and treatment | 0.05 (0.02) [0.02 to 0.08] | 0.05 (0.03) [0.00 to 0.14] | 0.03 (0.02) [0.00 to 0.09] |
| Comorbidity count | Trial | 0.07 (0.01) [0.05 to 0.09] |  |  |
| Comorbidity count | Trial and condition | 0.07 (0.01) [0.05 to 0.09] | 0.02 (0.02) [0.00 to 0.06] |  |
| Comorbidity count | Trial, condition and treatment | 0.04 (0.02) [0.02 to 0.07] | 0.03 (0.02) [0.00 to 0.08] | 0.05 (0.02) [0.00 to 0.10] |
| Asian | Trial | 0.07 (0.02) [0.04 to 0.11] |  |  |
| Asian | Trial and condition | 0.07 (0.02) [0.04 to 0.11] | 0.04 (0.03) [0.00 to 0.12] |  |
| Asian | Trial, condition and treatment | 0.04 (0.03) [0.00 to 0.10] | 0.04 (0.03) [0.00 to 0.13] | 0.06 (0.03) [0.01 to 0.13] |
| Black | Trial | 0.08 (0.02) [0.04 to 0.12] |  |  |
| Black | Trial and condition | 0.08 (0.02) [0.04 to 0.13] | 0.04 (0.03) [0.00 to 0.12] |  |
| Black | Trial, condition and treatment | 0.06 (0.03) [0.01 to 0.12] | 0.04 (0.03) [0.00 to 0.12] | 0.05 (0.03) [0.00 to 0.13] |
| Indigenous | Trial | 0.11 (0.05) [0.04 to 0.22] |  |  |
| Indigenous | Trial and condition | 0.11 (0.05) [0.05 to 0.23] | 0.08 (0.09) [0.00 to 0.31] |  |
| Indigenous | Trial, condition and treatment | 0.08 (0.06) [0.00 to 0.22] | 0.09 (0.09) [0.00 to 0.32] | 0.09 (0.06) [0.00 to 0.25] |
| Other | Trial | 0.16 (0.04) [0.09 to 0.24] |  |  |
| Other | Trial and condition | 0.14 (0.04) [0.08 to 0.23] | 0.08 (0.07) [0.00 to 0.24] |  |
| Other | Trial, condition and treatment | 0.11 (0.05) [0.04 to 0.21] | 0.07 (0.07) [0.00 to 0.25] | 0.11 (0.06) [0.01 to 0.24] |

**Figure 3.**
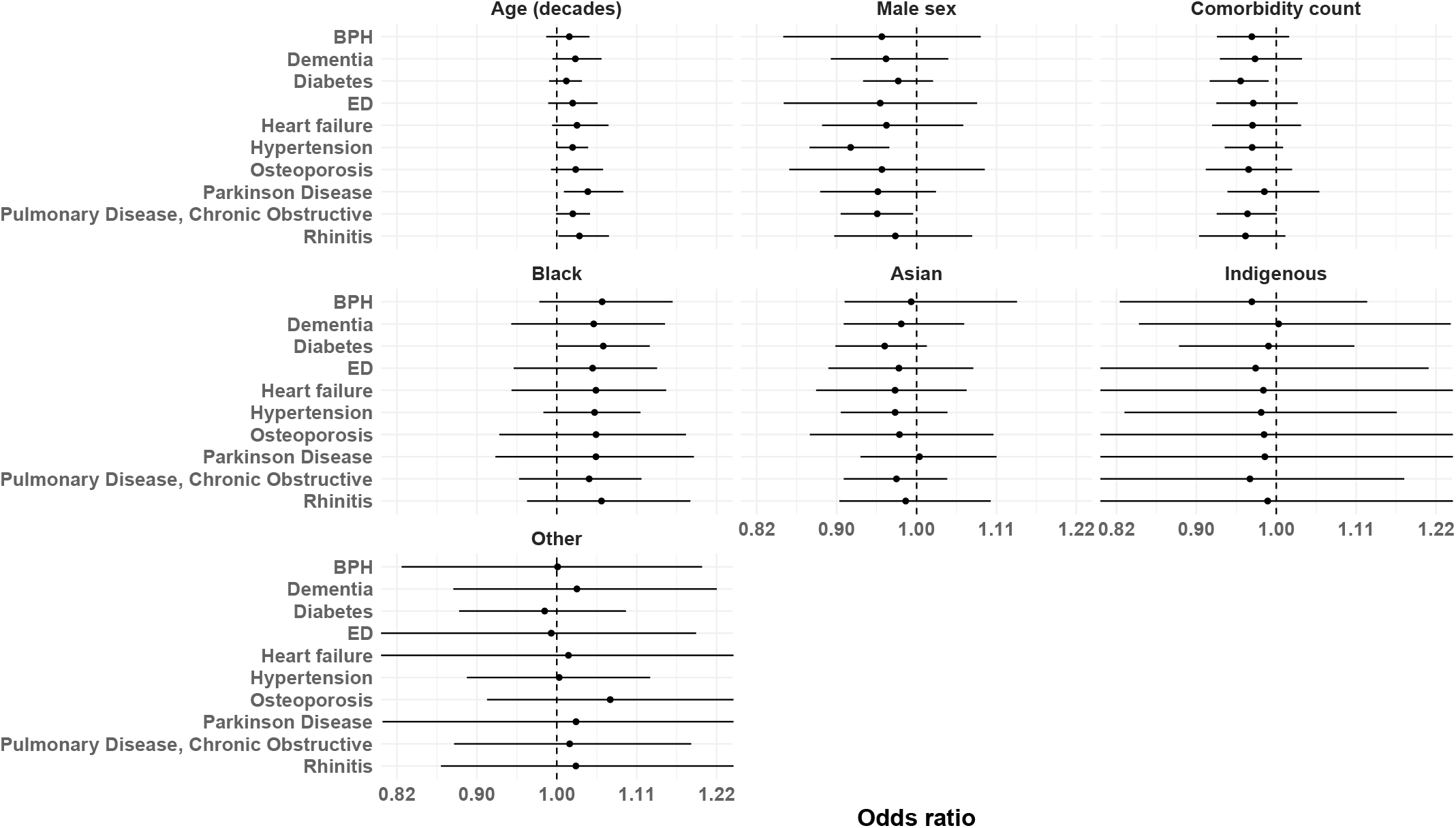
Forest plots showing mean odds ratio (OR) and 95% credible intervals of likelihood of screen failure for any reason by index condition. Results are displayed for age (per 10-year increase), sex (male versus female), comorbidity count (per 1 additional comorbidity) and self-reported /ethnicity. The black dotted line is the reference line (no effect OR = 1).

**Figure 4.**
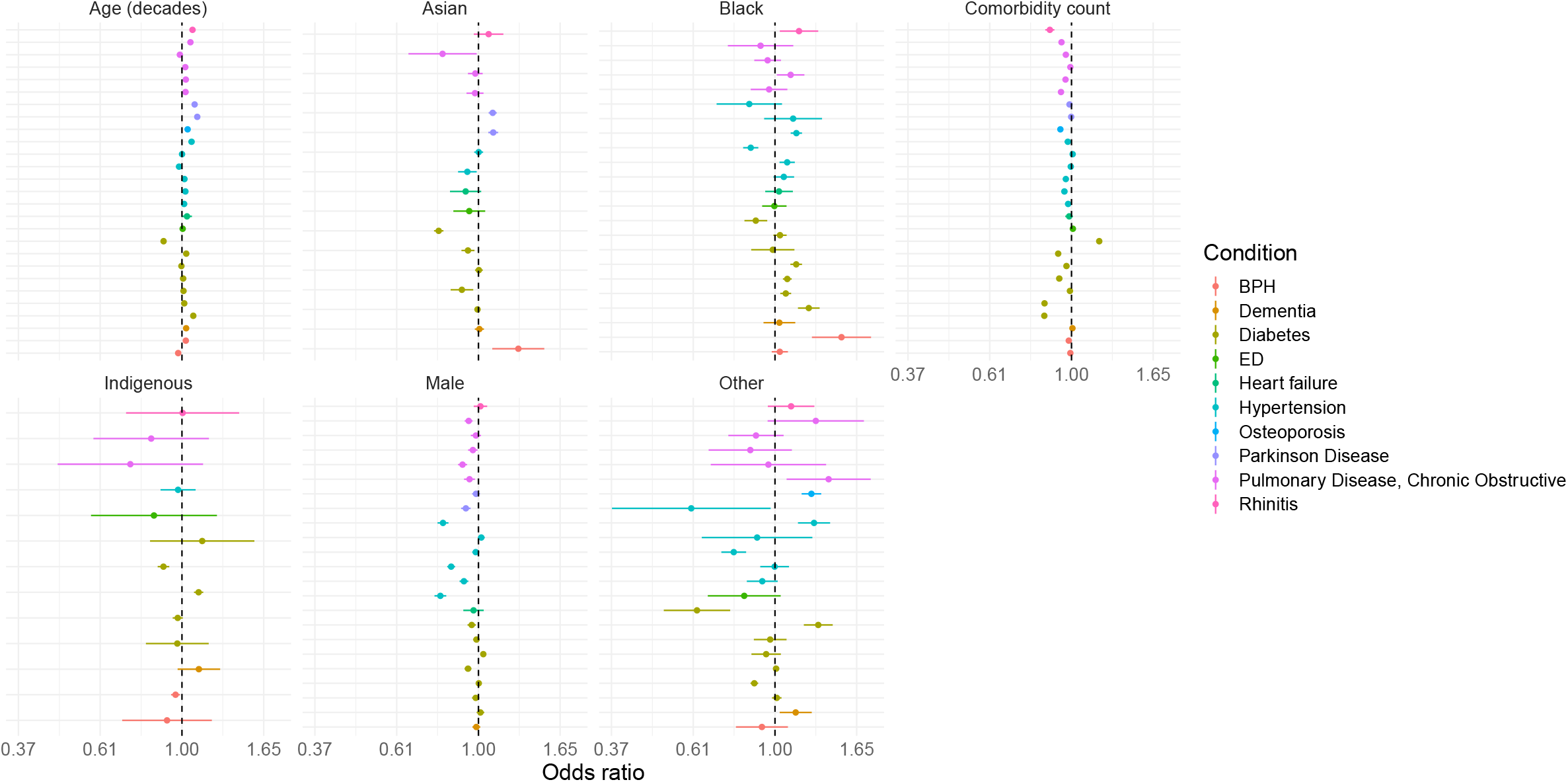
Forest plots showing mean odds ratio (OR) and 95% credible intervals of likelihood of screen failure for any reason for individual trials. Results are displayed for age (per 10-year increase), sex (male versus female), comorbidity count (per 1 additional comorbidity) and self-reported race/ethnicity. The black dotted line is the reference line (no effect OR = 1).

### Model diagnostics

The analysis code, model outputs from the trial-level logistic regression models fit within the trial safe havens, and the model outputs from the Bayesian hierarchical models, are available in the project GitHub repository (https://github.com/ChronicDiseaseEpi/screenfail_public). For the latter we also provide model diagnostics in terms of the number of divergent transitions, the Rhat and the bulk and tail effective sample sizes. There were no divergent transitions for any of the models. Rhat (a convergence diagnostic which compares the between-chain and within-chain estimates) was always <= 1.02 and for most models and terms was < 1.01, indicating satisfactory convergence. For some of the models where index condition was ignored, and those where trial was nested within index condition and treatment comparison the effective sample size was less than 400. However, for all the models presented in the main manuscript the effective sample sizes (bulk and tail) were >400.

## Discussion

Aggregate data analyses of randomised trial participants show underrepresentation of women, those with multi-morbidity and non-white ethnic groups^1–3^. However, it is not clear whether this under-representation arises at the invitation or screening phase. In this IPD analysis of 52 trials in chronic medical conditions, there was a weak association between higher age and increased likelihood of screen failure, with a lower likelihood of screen failure in individuals of male sex, although the credible interval for the latter included the null. Considering the detected associations between participant characteristics and screen failure were small, under-representation may be more driven by selection at the invitation to screening phase, rather than by application of trial eligibility criteria by the trial team during screening.

### Strengths and limitations

The strengths of this study are in the use of IPD across diverse trials conducted in chronic medical conditions, while limited previous comparisons have used aggregate trial data, questionnaires or been limited to particular index conditions. However, we acknowledge some limitations. Firstly, data describing screened populations are not routinely reported either in clinical trial repositories (such as ClinicalTrials.gov) nor in published trials: we cannot be certain of the accuracy of data collection within potential trial participants who have not consented to complete data collection^19^. Nevertheless, this limitation also demonstrates the scarcity and value of our data. Secondly, there were inadequate data available within the IPD to which we had access to allow us to examine the reasons for failing screening. While underlying associations for screen failure were weak overall, specific reasons for failing screening may be for other reasons, such as frailty, which we have not investigated. Thirdly, reflecting our initial trial selection, trials conducted in cancer, infections, psychiatric and developmental disorders were excluded. Similarly, we were only able to obtain IPD for trials contained within the Vivli trial data-sharing repository, and for sponsors who share data using this repository. It is possible that these findings are not representative of trials. Due to incomplete reporting of data on screen failures in trial registries such as ClinicalTrials.gov, we cannot measure the representativeness of these included trials for assessment of screen failure across other disease groups, sponsors or in non-industry funded trials; however, we previously illustrated that IPD trials are broadly representative of trials registered on ClinicalTrials.gov for assessment of trial attrition^16^. Finally, our measure of comorbidity was crude – an overall count, and it is plausible that either specific comorbidities, interactions between comorbidities, and/or interactions between comorbidities and the index condition are predictive of screen failure.

### Comparison with the previous literature

This is the first exploration examining participant characteristics associated with failing screening using trial IPD, across a wide variety of phase 3 and 4 industry-funded trials conducted for chronic medical conditions. However, a few studies have examined trial selection using other methodologies. A nationally representative survey found that women and men were equally likely to be invited to participate in trials^3^. We demonstrate that women are slightly more likely to fail trial screening than men across most index conditions, and clearly more likely to fail screening in trials of hypertension and chronic obstructive pulmonary disease. In a previous analysis of trial IPD, we showed that attrition after randomisation is not more likely in women^16^. Together, these studies suggest that enhancing the proportion of women invited to screening may improve female representation in trials.

We identified a weak and inconsistent association between Black race/ethnicity and increased likelihood of failing screening. Our findings are in keeping with the literature, which shows that people from racial and ethnic minorities are not substantially more likely to decline trial participation if offered^20,21^, but remain systematically under-represented in trials ^22–24^. This has prompted the development of guidelines to recruit and retain participants from ethnic minority groups (Trial Forge Guidance 3)^2^. The guidelines point to unintended exclusions of ethnic minorities because of restrictive eligibility criteria and recruitment pathways (certain comorbidities are more common among ethnic minorities^25,26^); provision of trial materials and information in poorly accessible forms (e.g., failure to consider language support, differences in literacy or cultural differences in the nature of communication); lack of cultural competence among trial staff; and an absence of trusting relationships between trialists and ethnic minority people. Ethnic minority groups may also have different motivations for trial participation, particularly in countries where universal healthcare is not provided^3^, and may stem from historical events (e.g., Tuskegee syphilis study), as well as discrimination that persists^27^. Since we found at most a weak association between Black race/ethnicity and screen failure among screened participants, this suggests that under-representation is more likely to have arisen at the invitation rather than the screening phase. Furthermore, our findings show important heterogeneity in patterns across groups, highlighting the importance of studying specific racial and ethnic groups. Consequently, approaches to improve representation may also be more effective if targeted at the invitation phase.

We identified a paradoxical association such that lower comorbidity count was associated with increased likelihood of failing screening; however, in sensitivity analyses – excluding people with very low levels of comorbidity – there was no apparent association between comorbidity count and screen failure. The most likely explanation for this observation is reporting or recording bias: potential participants may be more likely to recall medications and conditions when they have decided to participate in a trial, or investigators may make greater efforts to record such information in individuals they think are unlikely to fail screening.

### Implications for practice and policy

Ours is the first of which we are aware to meta-analyse associations between individual-level characteristics and failing to pass trial screening, and it was only possible due to our access to trial individual-level participant data. To better understand and improve trial representativeness, reporting guideline groups (such as CONSORT), representatives of journals (such as the International Committee of Medical Journal Editors), and trial registries which mandate results reporting (such as ClincialTrials.gov) may wish to consider requiring reporting of invited and screened participants as part of trial dissemination.

In lieu of more widespread reporting, our own findings – while limited to a relatively small and selected set of phase 3 and 4 industry-funded trials for which IPD were available – suggest that processes during the invitation to screening phase may be more important as regards trial representativeness.

## Conclusion

Age, sex, comorbidity count and race/ethnicity were not strongly associated with increased likelihood of screen failure. Proportionate increases in screening these underserved populations may improve representation in trials.

## Data Availability

Individual patient-level data are available from the Vivli Centre for Global Clinical Research Data platform (https://vivli.org). Trial-level results, model outputs and analysis code are provided on the project GitHub repository: https://github.com/ChronicDiseaseEpi/screenfail_public

https://github.com/ChronicDiseaseEpi/screenfail_public

## Contributorship statement

D.A.M. conceived the idea for the article. P.H. and E.B. identified suitable trials for inclusion. D.A.M., S.H.W., F.S.M., B.G., N.W. and S.D. critically advised on statistical analysis and presentation. J.S.L., J.C. and D.A.M. carried out the analysis. J.S.L. and D.A.M. created tables and figures. J.S.L. wrote the first draft of the manuscript. K.G. and S.V.K. critically advised on presentation and interpretation. J.S.L. and D.A.M. are guarantors for the overall content. All authors reviewed and approved the final submitted manuscript.

## Transparency declaration

The lead author affirms that the manuscript is an honest, accurate and transparent account of the study being reported.

## Public and Patient Involvement statement

Patients and the public were involved directly in the trial data used for this analysis, but were not directly involved in the design or planning of the current analysis. This paper has important messages for patients and the public, and support from Patient and Public Involvement and Engagement (PPIE) groups at the University of Glasgow will be sought to assist with dissemination of the research message.

## Ethical approval

Ethical approval was not required for this study.

## Declaration

This manuscript is based in part on research using data from data contributors, Boehringer Ingelheim, Eli Lilly, Roche, Takeda and UCB, that has been made available through Vivli, Inc. Vivli has not contributed to or approved, and Vivli, Boehringer Ingelheim, Eli Lilly, Roche, Takeda and UCB, are not in any way responsible for, the contents of this manuscript. SVK acknowledges funding from the Medical Research Council (MC_UU_00022/2) and the Scottish Government Chief Scientist Office (SPHSU17). Outside the submitted work, JSL acknowledges personal lectureship honoraria from Astra Zeneca, Pfizer and Bristol Myers Squibb.

## Data sharing statement

Individual patient-level data are available from the Vivli Centre for Global Clinical Research Data platform (https://vivli.org). Trial-level results, model outputs and analysis code are provided on the project GitHub repository:

- https://github.com/ChronicDiseaseEpi/screenfail_public

## Supplementary Data file

**Table S1.** Trial level models for the mean odds ratio (standard error) [95% credible interval] for screen failure. Model A: adjusted for age, sex and age:sex interaction; Model B: adjusted for age, sex, comorbidity count and age:sex:comorbidity count interaction; Model C: adjusted for age, sex, comorbidity count, race/ethnicity and sex:race/ethnicity interaction. For all models, trial was nested within condition, then condition and treatment.

| Coefficient | Model | Model A<br>52 trials | Model B<br>31 trials | Model C<br>27 trials |
| --- | --- | --- | --- | --- |
| Age (decades) | Trial and condition | 1.00 (0.98 to 1.03) | 1.02 (0.99 to 1.05) | 1.02 (1.00 to 1.04) |
| Age (decades) | Trial, condition and treatment | 1.00 (0.98 to 1.03) | 1.02 (0.99 to 1.05) | 1.02 (1.00 to 1.04) |
| Male | Trial and condition | 0.98 (0.90 to 1.08) | 0.97 (0.80 to 1.14) | 0.96 (0.91 to 1.03) |
| Male | Trial, condition and treatment | 0.98 (0.90 to 1.08) | 0.97 (0.79 to 1.16) | 0.97 (0.90 to 1.05) |
| Comorbidity count | Trial and condition |  | 0.97 (0.91 to 1.04) | 0.97 (0.94 to 1.00) |
| Comorbidity count | Trial, condition and treatment |  | 0.98 (0.91 to 1.05) | 0.97 (0.94 to 1.02) |
| Asian | Trial and condition |  |  | 1.00 (0.93 to 1.08) |
| Asian | Trial, condition and treatment |  |  | 1.00 (0.92 to 1.09) |
| Black | Trial and condition |  |  | 1.09 (1.00 to 1.19) |
| Black | Trial, condition and treatment |  |  | 1.08 (0.99 to 1.20) |
| Indigenous | Trial and condition |  |  | 1.00 (0.84 to 1.20) |
| Indigenous | Trial, condition and treatment |  |  | 1.01 (0.83 to 1.24) |
| Other | Trial and condition |  |  | 1.01 (0.88 to 1.15) |
| Other | Trial, condition and treatment |  |  | 1.01 (0.87 to 1.17) |
| Interaction male:Asian | Trial and condition |  |  | 0.97 (0.91 to 1.03) |
| Interaction male:Asian | Trial, condition and treatment |  |  | 0.97 (0.91 to 1.05) |
| Interaction male:Black | Trial and condition |  |  | 0.90 (0.70 to 1.08) |
| Interaction male:Black | Trial, condition and treatment |  |  | 0.91 (0.71 to 1.12) |
| Interaction male: Indigenous | Trial and condition |  |  | 0.90 (0.38 to 2.03) |
| Interaction male: Indigenous | Trial, condition and treatment |  |  | 0.88 (0.35 to 2.14) |
| Interaction male:Other | Trial and condition |  |  | 1.00 (0.75 to 1.34) |
| Interaction male:Other | Trial, condition and treatment |  |  | 1.00 (0.74 to 1.32) |
| Interaction male:age | Trial and condition | 1.00 (0.98 to 1.01) | 1.00 (0.97 to 1.03) |  |
| Interaction male:age | Trial, condition and treatment | 1.00 (0.98 to 1.01) | 1.00 (0.96 to 1.04) |  |
| Interaction male:comorbidity count | Trial and condition |  | 0.96 (0.86 to 1.07) |  |
| Interaction male:comorbidity count | Trial, condition and treatment |  | 0.95 (0.84 to 1.08) |  |
| Interaction age:comorbidity count | Trial and condition |  | 1.00 (0.99 to 1.01) |  |
| Interaction age:comorbidity count | Trial, condition and treatment |  | 1.00 (0.99 to 1.01) |  |
| Interaction age:male:comorbidity count | Trial and condition |  | 1.01 (0.99 to 1.03) |  |
| Interaction age:male:comorbidity count | Trial, condition and treatment |  | 1.01 (0.99 to 1.03) |  |

**Table S2.** Models for the log odds ratio (standard error) [95% credible interval] for screen failure examining variation in estimates between trial, between condition and between trial and condition. Model 1: adjusted for age and sex; Model 2: comorbidity count only; Model 3: race/ethnicity only; Model 4: adjusted for age, sex and comorbidity count; Model 5: adjusted for age, sex, comorbidity count and race/ethnicity. Models were conducted at three levels: trial (where condition and treatment were ignored); trial nested within condition; and trial nested within condition and treatment.

| Coefficient | Model | Model 1 - 52 trials | Model 2 - 31 trials | Model 3 - 45 trials | Model 4 - 31 trials | Model 5 - 27 trials |
| --- | --- | --- | --- | --- | --- | --- |
| Age (decades) | Trial |  |  |  |  | 1.02 (1.00 to 1.03) |
| Age (decades) | Trial and condition | 1.01 (0.99 to 1.03) |  |  | 1.02 (1.01 to 1.04) | 1.02 (1.00 to 1.04) |
| Age (decades) | Trial, condition and treatment | 1.01 (0.99 to 1.03) |  |  | 1.02 (1.00 to 1.04) | 1.02 (1.00 to 1.04) |
| Male | Trial |  |  |  |  | 0.95 (0.93 to 0.98) |
| Male | Trial and condition | 0.97 (0.93 to 1.01) |  |  | 0.95 (0.91 to 1.00) | 0.96 (0.91 to 1.01) |
| Male | Trial, condition and treatment | 0.97 (0.93 to 1.01) |  |  | 0.95 (0.91 to 1.00) | 0.96 (0.91 to 1.02) |
| Comorbidity count | Trial |  |  |  |  | 0.97 (0.94 to 0.99) |
| Comorbidity count | Trial and condition |  | 0.97 (0.95 to 1.00) |  | 0.97 (0.94 to 1.00) | 0.97 (0.94 to 1.00) |
| Comorbidity count | Trial, condition and treatment |  | 0.98 (0.94 to 1.01) |  | 0.98 (0.94 to 1.02) | 0.98 (0.94 to 1.02) |
| Asian | Trial |  |  |  |  | 0.97 (0.93 to 1.01) |
| Asian | Trial and condition |  |  | 0.98 (0.93 to 1.04) |  | 0.98 (0.93 to 1.04) |
| Asian | Trial, condition and treatment |  |  | 0.98 (0.92 to 1.05) |  | 0.98 (0.91 to 1.05) |
| Black | Trial |  |  |  |  | 1.05 (1.00 to 1.10) |
| Black | Trial and condition |  |  | 1.04 (0.99 to 1.09) |  | 1.05 (0.98 to 1.12) |
| Black | Trial, condition and treatment |  |  | 1.04 (0.99 to 1.09) |  | 1.05 (0.97 to 1.12) |
| Indigenous | Trial |  |  |  |  | 0.98 (0.89 to 1.08) |
| Indigenous | Trial and condition |  |  | 0.94 (0.87 to 1.03) |  | 0.98 (0.84 to 1.15) |
| Indigenous | Trial, condition and treatment |  |  | 0.95 (0.87 to 1.05) |  | 0.98 (0.82 to 1.16) |
| Other | Trial |  |  |  |  | 1.01 (0.92 to 1.09) |
| Other | Trial and condition |  |  | 1.01 (0.93 to 1.10) |  | 1.02 (0.91 to 1.15) |
| Other | Trial, condition and treatment |  |  | 1.02 (0.94 to 1.12) |  | 1.02 (0.90 to 1.16) |

